# Bayesian Genome-wide Polygenic Score Integration with FRAX for Enhanced Fracture Risk Prediction in Postmenopausal Women

**DOI:** 10.1101/2025.06.06.25329139

**Authors:** Anqi Liu, Jianing Liu, Qing Wu

## Abstract

**Importance:** Current fracture risk prediction tools, including the Fracture Risk Assessment Tool (FRAX), do not incorporate genetic risk factors limiting accuracy and contributing to misclassification and suboptimal care.

**Objective:** To develop and validate a novel genome-informed fracture risk assessment tool (Bayes-FRAX) integrating Bayesian genome-wide polygenic scores (GPS) into the established FRAX to enhance major osteoporotic fracture (MOF) prediction.

**Design, Setting, and Participants:** This retrospective cohort study analyzed clinical and genetic data from 6,932 postmenopausal women enrolled in the Women’s Health Initiative (1993-1998).

**Exposures:** Integration of GPS derived using Polygenic Risk Score Continuous Shrinkage (PRS-CS) and Summary data-based Bayesian regression (SBayesR) into FRAX.

**Main Outcomes and Measures:** Primary outcomes included the incidence of MOF. Predictive performance metrics assessed were area under the receiver operating characteristic curve (AUROC), area under the precision-recall curve (AUPRC), calibration slopes, Hosmer–Lemeshow goodness-of-fit tests, net reclassification improvement (NRI), diagnostic sensitivity, decision curve analysis (DCA), and external validation metrics.

**Results:** Of 6,932 women, 513 (7.4%) experienced MOF. Bayes-FRAX significantly improved prediction over standard FRAX based solely on clinical risk factors, increasing AUROC from 0.662 to 0.680 for both PRS-CS and SBayesR. AUPRC improved from 0.120 (FRAX) to 0.140 (PRS-CS) and 0.138 (SBayesR). Calibration slopes were ideal (GPS-PRS-CS: 1.00 [95% CI: 0.8569–1.1431]; GPS-SBayesR: 1.00 [95% CI: 0.8560–1.1437]). Bayes-FRAX reclassified 3.5% of women, 34% near the intervention threshold. NRI improved by 4.59% (SBayesR) and 4.34% (PRS-CS), largely from better classification of women who fractured (5.85% and 5.65%). Decision curve analyses demonstrated greater net clinical benefit at clinically relevant thresholds, notably at the 20% threshold. External validation in 852 independent White postmenopausal women confirmed robust generalizability, with GPS significantly associated with fracture risk (PRS-CS OR = 0.148, 95% CI: 0.052–0.411; SBayesR OR = 0.116, 95% CI: 0.040–0.324). Likelihood ratio tests also supported improved model fit after GPS inclusion (PRS-CS: *P* < 0.001; SBayesR: *P* <0.001). Sensitivity analysis without BMD demonstrated stable AUROC (0.74).

**Conclusions and Relevance:** Integrating GPS into FRAX using Bayesian methods improved fracture risk prediction, reclassification, and decision-making. Bayes-FRAX provides a generalizable tool for personalized osteoporosis care, especially for women near treatment thresholds.

**Key Points:** *Question:* Does incorporating genome-wide polygenic scores using Bayesian methods improve fracture risk prediction beyond the traditional FRAX?

*Findings:* In this cohort study of 6,932 postmenopausal women, adding Bayesian polygenic scores significantly improved prediction accuracy, calibration, and clinical reclassification, particularly among women older than 65 years, with robust external validation.

*Meaning:* Bayes-FRAX provides a personalized, more accurate fracture risk assessment, potentially improving clinical decision-making for postmenopausal women near treatment thresholds.

## Introduction

Osteoporosis, characterized by reduced bone density and compromised skeletal microarchitecture, poses a growing global health challenge due to aging populations and rising incidences of fragility fractures. In the United States alone, approximately 10 million adults older than 50 years have osteoporosis, and an additional 34 million are at elevated risk, collectively resulting in nearly 1.5 million osteoporotic fractures annually[1]. These fractures substantially increase morbidity, mortality, healthcare expenditures, and profoundly impair patient quality of life[2]. Enhancing the accuracy of fracture prediction tools to facilitate precise clinical intervention remains an urgent public health priority.

Currently, fracture risk prediction heavily relies on clinical risk factors, including low bone mineral density (BMD), advanced age, low body mass index (BMI), a history of previous fractures, rheumatoid arthritis, glucocorticoid use, and a family history of fractures[3]. The Fracture Risk Assessment Tool (FRAX), a widely adopted clinical algorithm developed by the University of Sheffield in collaboration with the WHO Collaborating Centre for Metabolic Bone Diseases, estimates the 10-year fracture probability based on these clinical factors[4, 5]. Despite widespread clinical adoption, FRAX demonstrates only modest accuracy in predicting major osteoporotic fractures among postmenopausal women, with reported AUROC values commonly ranging from approximately 0.55 to 0.65, indicating substantial limitations in fracture risk stratification and highlighting the need for improvement.[6, 7]. Recent studies have highlighted FRAX’s limitations, particularly in accurately identifying individuals near critical treatment thresholds, which may lead to suboptimal preventive interventions and uncertainty in clinical decision-making.[8]

Genetic studies have consistently demonstrated substantial heritability of osteoporosis[9], with genetic factors accounting for approximately 50% to 80% of variability in BMD and fracture risk[10–14]. Genome-wide association studies (GWAS) have identified numerous single-nucleotide polymorphisms (SNPs) associated with osteoporosis-related traits[15–17]. However, traditional polygenic risk scores (PRS), derived primarily from genome-wide significant SNPs, have demonstrated limited incremental predictive value beyond conventional clinical factors, capturing only a fraction of the underlying genetic susceptibility[18–20]. In contrast, genome-wide polygenic scores (GPS), aggregating the effects of many more genetic variants across the genome, have shown significantly enhanced predictive performance[21–23].

Despite these genetic advancements, current clinical fracture prediction models, including FRAX, do not yet incorporate genetically informed risk scores, representing a significant translational gap. Bayesian polygenic methodologies have recently emerged as particularly promising approaches[24, 25], explicitly accounting for linkage disequilibrium and capturing the broader polygenic architecture of complex traits like osteoporosis. By addressing these genetic complexities, Bayesian methods offer the potential for improved predictive accuracy and calibration compared with traditional genetic approaches, thereby directly addressing critical limitations identified in conventional fracture prediction tools.

To address this critical gap explicitly, we developed and validated Bayes-FRAX, the first fracture risk assessment tool integrating Bayesian-derived genome-wide polygenic scores with established FRAX clinical parameters. Specifically targeting postmenopausal women—a population particularly vulnerable to rapid bone loss and heightened fracture risk—this study evaluates whether incorporating Bayesian-derived genetic scores meaningfully improves fracture prediction accuracy, clinical reclassification, and treatment eligibility classification. Improved fracture risk prediction through Bayes-FRAX has the potential to substantially enhance personalized clinical decision-making, optimize preventive interventions, and ultimately improve patient outcomes in osteoporosis management.

## Materials and Methods

### 2.1 Study Design and Population

This retrospective cohort study utilized clinical and genetic data from postmenopausal women enrolled in the Women’s Health Initiative (WHI), specifically participants from the Genomics and Randomized Trials Network (GARNET), WHI Memory Study (WHIMS), and SNP Health Association Resource (SHARe). Data was accessed via the Database of Genotypes and Phenotypes (dbGaP; accession number phs000200.v12. p3). Participants were eligible if genetic data, complete clinical variables required for FRAX calculation, and documented follow-up fracture outcomes were available. Explicit exclusion criteria included participants with >20% missing clinical data and missing fracture outcome data. The final analytic sample comprised 6,932 postmenopausal women. (Figure 1)

**Figure 1.**
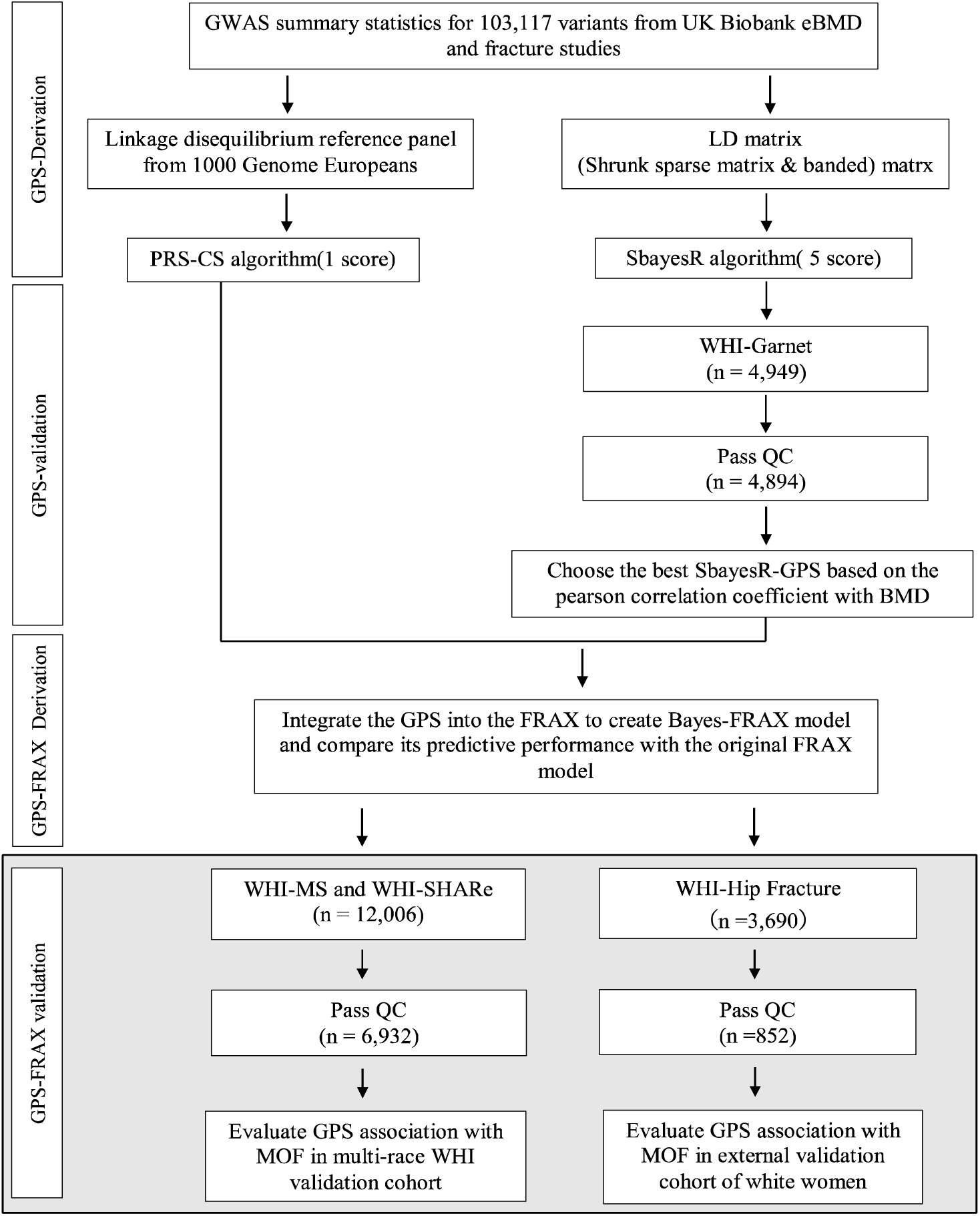
Workflow for Derivation, Validation, and Integration of Genome-wide Polygenic Scores (GPS). GPS was derived from 103,117 genetic variants obtained from the Genetic Factors for Osteoporosis Consortium (GEFOS). Polygenic Risk Score Continuous Shrinkage (PRS-CS) generated one score; Summary-data-based Bayesian Regression (SBayesR) generated five scores using multiple linkage disequilibrium (LO) structures and Bayesian priors. The best-performing GPS was identified based on the highest Pearson correlation with bone mineral density (BMD) in the Genomics and Randomized Trials Network (GARNET) sub-study (n=2,458). The selected GPS was validated in Women’s Health Initiative (WHI) sub-studies (n=6,932), and integrated into FRAX to create Bayes-FRAX for clinical fracture risk prediction. Detailed Bayesian parameters are provided in eMethod1. Abbreviations: GPS, Genome-wide Polygenic Score; SNP, Single-Nucleotide Polymorphism; GWAS, Genome-wide Association Study; PRS-CS, Polygenic Risk Score Continuous Shrinkage; SBayesR, Summary-data-based Bayesian Regression; GEFOS, Genetic Factors for Osteoporosis Consortium; BMD, Bone Mineral Density; GARNET, Genomics and Randomized Trials Network; WHI, Women’s Health Initiative; FRAX, Fracture Risk Assessment Tool; MOF, Major Osteoporotic Fracture.

### 2.2 Fracture Ascertainment and Clinical Data

Fracture outcomes encompassed MOF, defined as fractures of the hip, spine, wrist, or proximal humerus. Fractures were ascertained by participant self-report. While self-reported fracture history may be subject to recall bias, prior validation studies within the WHI cohort have demonstrated high reliability and specificity, particularly for hip and other types of MOF [26]. Clinical risk factors for FRAX calculation were collected at baseline, explicitly including age, body mass index (BMI), prior fracture history, rheumatoid arthritis, glucocorticoid use, parental hip fracture history, smoking status, secondary osteoporosis causes, and alcohol consumption (>3 drinks/day). BMD at the hip was assessed via dual-energy X-ray absorptiometry (DXA) in a subset; however, primary models utilized clinical risk factors alone.

### 2.3 Genotype Data and Quality Control

Genotyping was conducted using Illumina and Affymetrix platforms, with imputation using the 1000 Genomes Project (Phase 3 European reference panel)[27]. Rigorous quality control explicitly included sample call rates ≥95%, SNP call rates ≥90%, Hardy-Weinberg equilibrium (p ≥ 1 × 10⁻⁶), and minor allele frequency (MAF ≥0.01). Genotype QC was conducted using PLINK[28].

### 2.4 Genome-Wide Polygenic Score (GPS) Calculation

To calculate genome-based polygenic scores (GPS), we utilized publicly available summary statistics from the 2018 release of the GEnetic Factors for OSteoporosis (GEFOS) Consortium, based on genome-wide association analyses of estimated BMD (eBMD) from the UK Biobank[17]. Post-QC, 103,117 genome-wide significant SNPs (imputation INFO ≥0.8, p ≤ 5 × 10⁻⁸) were available for genome-wide polygenic score (GPS) calculation.

GPS were explicitly calculated using two Bayesian methods: Polygenic Risk Score Continuous Shrinkage (PRS-CS) and Summary-data-based Bayesian regression (SBayesR).

PRS-CS employs continuous shrinkage prior on SNP effect sizes, explicitly incorporating linkage disequilibrium (LD) structure from a 1000 Genomes European reference panel, robustly capturing fine-scale LD patterns and improving predictive accuracy[29].

SBayesR explicitly models SNP effects with a mixture of normal distributions, directly integrating LD structures through LD matrices. This method effectively addresses polygenic complexity and localized LD correlation via optimized hyperparameters[30, 31].

### 2.5 Bayes-FRAX Model Development

Bayes-FRAX explicitly integrated GPS into FRAX clinical risk factor (FRAX-CRF) models using logistic regression. The FRAX-predicted fracture probabilities (P) were logit-transformed, then adjusted with GPS, age, and GPS-age interaction explicitly:

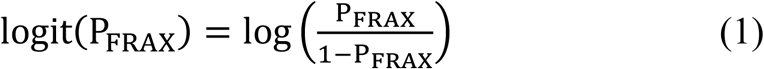

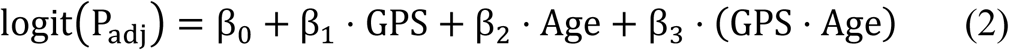

Probabilities were explicitly back-transformed using inverse logit:

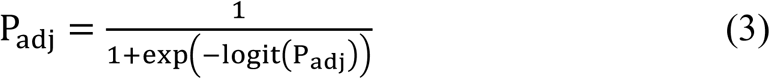

Separate models explicitly utilized GPS from PRS-CS and SBayesR.

### 2.6 Statistical Analysis

Baseline characteristics were summarized using means (standard deviation, SD) or frequencies (percentages). Between-group comparisons were explicitly performed using chi-square, t-tests, or Fisher’s exact test as appropriate.

Explicit methods for calibration assessment included calibration slopes and Hosmer–Lemeshow goodness-of-fit tests. Predictive stability was validated through rigorous five-fold internal cross-validation, reporting the mean AUROC and AUPRC across folds.

Decision Curve Analysis (DCA) explicitly quantified the net clinical benefit of Bayes-FRAX models compared to standard FRAX, assessing clinical utility across fracture probability thresholds (10%-30%). Diagnostic accuracy was evaluated against observed outcomes using McNemar’s tests, explicitly defining correct classification above or below clinical thresholds.

Treatment eligibility-based reclassification was performed using two guideline-based thresholds. A fixed 20% threshold was applied based on the U.S. National Osteoporosis Foundation (NOF)[32] and Age-dependent thresholds from the UK National Osteoporosis Guideline Group (NOGG)[33] were also evaluated. Reclassification rates and Net reclassification improvement (NRI) were computed to quantify gains in classification accuracy under each criterion.

### 2.7 Sensitivity and Subgroup Analyses

Explicit subgroup analyses stratified participants by age (≤65 vs. >65 years). Sensitivity analyses compared Bayes-FRAX performance with and without hip BMD data, confirming the robustness of findings.

### 2.8 External Validation

External validation was performed in an independent cohort of 852 White postmenopausal women (WHI-Hip Fracture, a sub-study of the Women’s Health Initiative). Predictive utility was evaluated by assessing the association between GPS and fracture risk using odds ratios and by testing model fit improvement through likelihood ratio tests.

### 2.9 Software and Statistical Packages

All analyses utilize R software version 4.2.2 (R Foundation for Statistical Computing)[34]. We employed packages including “pROC” (ROC analyses), “PredictABEL” (reclassification analyses), and “rmda” (decision curve analyses) for this study.

### 2.10 Ethical Considerations and Data Availability

All analyses were conducted under Institutional Review Board approval from Ohio State University (IRB approval number 2022H0420). Genotype and phenotype data are available via dbGaP under approved access agreements.

## Results

### 3.1 Study Population Characteristics

The final analytic cohort included 6,932 postmenopausal women with a mean (SD) age of 64.2 (7.3) years. 513 participants (7.4%) experienced a MOF. Compared to participants without fractures, those with fractures were significantly older (mean [SD], 68.3 [6.4] vs. 63.9 [7.3] years; P<0.01), had lower body weight (BMI; mean [SD], 73.2 [15.0] vs. 77.7 [16.4] kg; P<0.01), lower hip bone mineral density (BMD; mean [SD], 0.79 [0.13] vs. 0.89 [0.15] g/cm²; P<0.01), and higher baseline FRAX-predicted 10-year MOF risk (mean [SD], 12.0% [7.9] vs. 8.3% [7.0]; P<0.01) (Table 1).

**Table 1.**
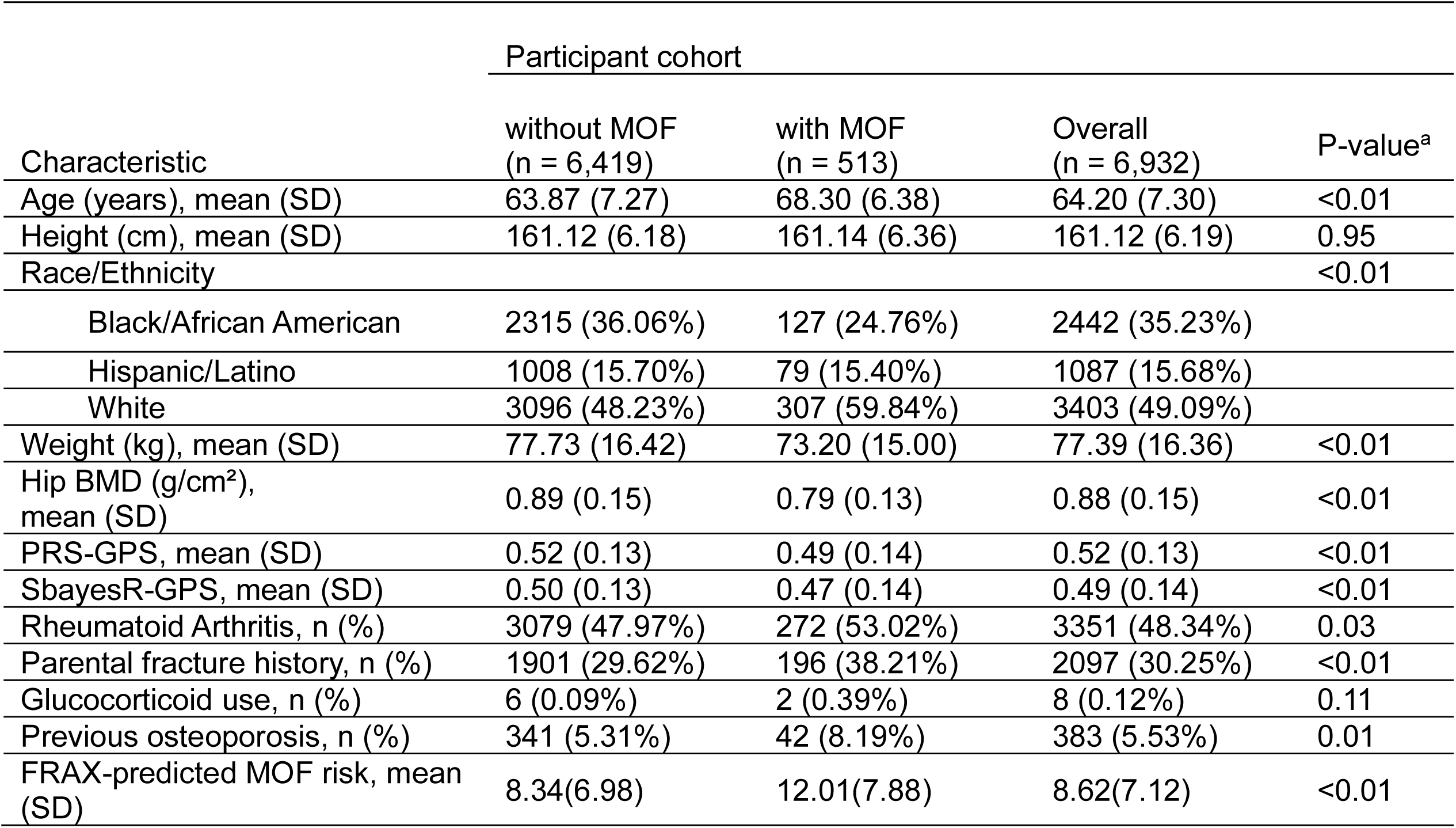
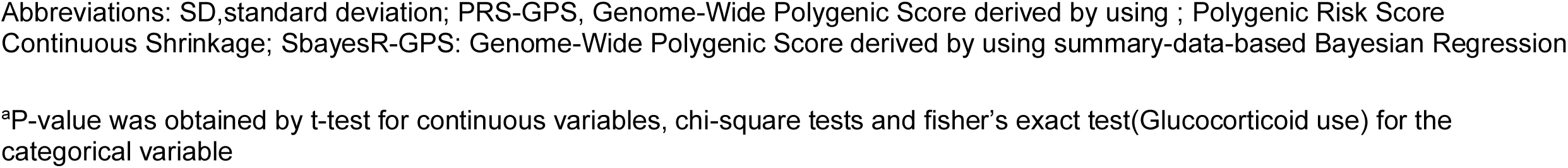
Baseline characteristics of 6,932 women in the WHI independent testing cohort, stratified by major osteoporotic fracture (MOF) status.

| Characteristic | Participant cohort |  |  | P-value<sup>a</sup> |
| --- | --- | --- | --- | --- |
|  | without MOF<br>(n = 6,419) | with MOF<br>(n = 513) | Overall<br>(n = 6,932) |  |
| Age (years), mean (SD) | 63.87 (7.27) | 68.30 (6.38) | 64.20 (7.30) | <0.01 |
| Height (cm), mean (SD) | 161.12 (6.18) | 161.14 (6.36) | 161.12 (6.19) | 0.95 |
| Race/Ethnicity |  |  |  | <0.01 |
| Black/African American | 2315 (36.06%) | 127 (24.76%) | 2442 (35.23%) |  |
| Hispanic/Latino | 1008 (15.70%) | 79 (15.40%) | 1087 (15.68%) |  |
| White | 3096 (48.23%) | 307 (59.84%) | 3403 (49.09%) |  |
| Weight (kg), mean (SD) | 77.73 (16.42) | 73.20 (15.00) | 77.39 (16.36) | <0.01 |
| Hip BMD (g/cm<sup>2</sup>), mean (SD) | 0.89 (0.15) | 0.79 (0.13) | 0.88 (0.15) | <0.01 |
| PRS-GPS, mean (SD) | 0.52 (0.13) | 0.49 (0.14) | 0.52 (0.13) | <0.01 |
| SbayesR-GPS, mean (SD) | 0.50 (0.13) | 0.47 (0.14) | 0.49 (0.14) | <0.01 |
| Rheumatoid Arthritis, n (%) | 3079 (47.97%) | 272 (53.02%) | 3351 (48.34%) | 0.03 |
| Parental fracture history, n (%) | 1901 (29.62%) | 196 (38.21%) | 2097 (30.25%) | <0.01 |
| Glucocorticoid use, n (%) | 6 (0.09%) | 2 (0.39%) | 8 (0.12%) | 0.11 |
| Previous osteoporosis, n (%) | 341 (5.31%) | 42 (8.19%) | 383 (5.53%) | 0.01 |
| FRAX-predicted MOF risk, mean (SD) | 8.34(6.98) | 12.01(7.88) | 8.62(7.12) | <0.01 |

Abbreviations: SD, standard deviation; PRS-GPS, Genome-Wide Polygenic Score derived by using ; Polygenic Risk Score Continuous Shrinkage; SbayesR-GPS: Genome-Wide Polygenic Score derived by using summary-data-based Bayesian Regression
<sup>a</sup>P-value was obtained by t-test for continuous variables, chi-square tests and fisher's exact test(Glucocorticoid use) for the categorical variable

### 3.2 Predictive Performance and Calibration

Bayes-FRAX models significantly improved fracture prediction compared with the standard FRAX-CRF. The AUROC increased from 0.662 (FRAX-CRF) to 0.680 (Bayes-FRAX PRS-CS;95% CI: 0.657-0.704, P = 0.045) and 0.680 (Bayes-FRAX SBayesR; 95% CI: 0.657-0.703, P =0.047) (eTable 1, eFigure 1). The AUPRC improved from 0.120 (FRAX-CRF) to 0.140 (PRS-CS) and 0.138 (SBayesR). Calibration slopes for the Bayes-FRAX models were near 1 (PRS-CS: 1.00 [0.857–1.143]; SBayesR: 1.00 [0.856–1.144]). Calibration assessment indicated significant deviations from ideal calibration for all models based on Hosmer–Lemeshow tests. However, Bayes-FRAX models demonstrated notably improved calibration, evidenced by substantially higher Hosmer–Lemeshow P values (PRS-CS: P = .001; SBayesR: P = .001) compared with the FRAX-CRF model (P < .0001) (Figure 2, eTable1). Internal validation via 5-fold cross-validation revealed stable predictive performance, with consistent AUROC values across validation folds for both PRS-CS (AUROC, 0.681) and SBayesR (AUROC, 0.680) models, confirming the robustness of Bayes-FRAX (eTable 2).

**Figure 2.**
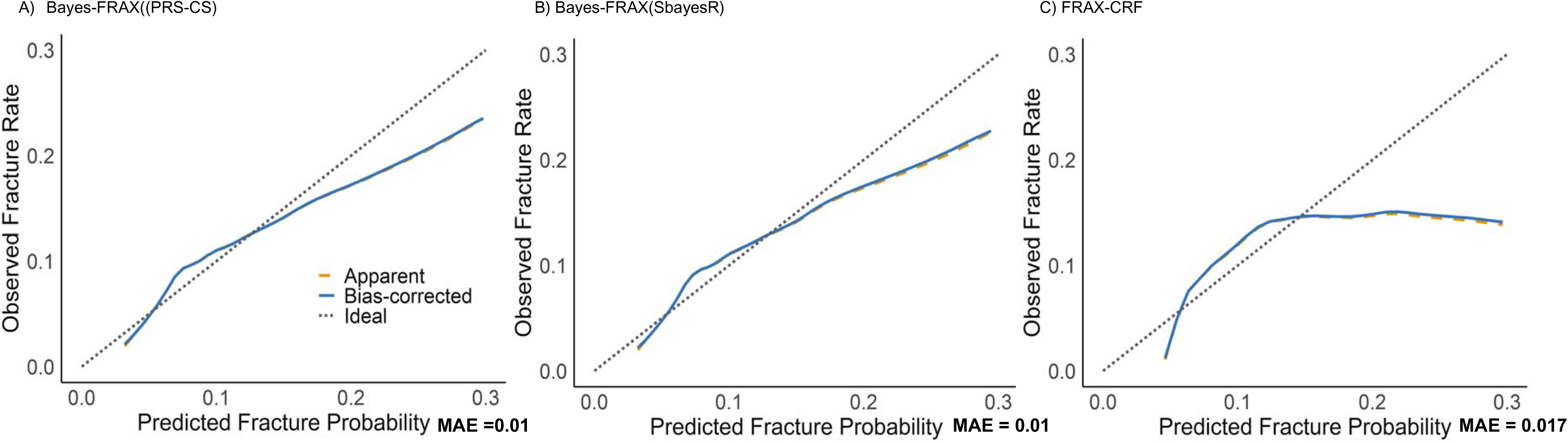
Calibration plots comparing Bayes-enhanced FRAX with standard FRAX. (A) Bayes-FRAX(PRS-CS) similarly shows strong calibration performance (MAE = 0.01). (B) Bayes-FRAX(SbayesR) demonstrates excellent calibration with very close alignment to the ideal diagonal line (MAE = 0.01). (C) FRAX-CRF without genetic data shows noticeable deviation from ideal calibration, particularly underestimating fracture probability at higher predicted risks (MAE = 0.017). Curves represent observed vs. predicted fracture probabilities, with bias-corrected calibration from 1,000 bootstrap repetitions. Diagonal dashed lines represent ideal calibration.

### 3.3 Clinical Reclassification and Net Reclassification Improvement (NRI)

At a fixed 20% treatment threshold for MOF, the Bayes-FRAX reclassified approximately 3.5% of women relative to FRAX-CRF (SBayesR: 3.48%; PRS-CS: 3.51%). Approximately 34% of these reclassified women had fracture risk estimates near the intervention threshold (15%-25%). NRI indicated overall gains of 4.59% (SBayesR) and 4.34% (PRS-CS), predominantly driven by correctly reclassifying women who subsequently experienced fractures (NRI for fracture events: 5.85% and 5.65%, respectively). Subgroup analyses demonstrated notably greater reclassification improvements among women older than 65 years (NRI: 5.04% [SBayesR]; 4.68% [PRS-CS]), whereas younger women (≤65 years) exhibited minimal NRI improvements (0.08%) (Table 2).

**Table 2.** Percentage reclassified and net reclassification improvement (NRI) for individual FRAX intervention criteria.

|  | Fixed MOF 20% | Age-dependent MOF (NOGG guideline) |
| --- | --- | --- |
| **Reclassification (%)** |  |  |
| all subjects (SbayesR) | 3.48% | 3.29% |
| all subjects (PRS-CS) | 3.51% | 3.30% |
| close to cut-off <sup>a</sup> SbayesR) | 33.92% | 6.47% |
| close to cut-off (PRS-CS) | 34.36% | 6.56% |
| age≤ 65 years SbayesR) | 0.08% | 0.19% |
| age≤ 65 years (PRS-CS) | 0.08% | 0.20% |
| age> 65 years (SbayesR) | 7.06% | 6.56% |
| age> 65 years (PRS-CS) | 7.12% | 6.59% |
| **Fracture outcome for NRI analysis** |  |  |
| NRI fracture, all subjects (SbayesR) | 5.85% | 5.46% |
| NRI fracture, all subjects (PRS-CS) | 5.65% | 5.46% |
| NRI non-fracture, all subjects (SbayesR) | -1.26% | -1.23% |
| NRI non-fracture, all subjects (PRS-CS) | -1.30% | -1.25% |
| NRI overall, all subjects (SbayesR) | 4.59% | 4.23% |
| NRI overall, all subjects (PRS-CS) | 4.34% | 4.21% |
| NRI overall, age 65 years (SbayesR) | 0.08% | -0.01% |
| NRI overall, age 65 years (PRS-CS) | 0.08% | -0.12% |
| NRI overall, age >65 years (SbayesR) | 5.04% | 4.62% |
| NRI overall, age >65 years (PRS-CS) | 4.68% | 4.58% |

<sup>a</sup> MOF±5%

### 3.4 Diagnostic Accuracy Using Observed Fracture Outcomes

To assess changes in clinical risk categorization, we compared Bayes-FRAX and FRAX-CRF classifications based on whether the predicted 10-year MOF risk exceeded the standard 20% treatment threshold. Bayes-FRAX (PRS-CS) reclassified 519 of 6,932 women (7.5%), including 51 (0.7%) from low to high risk (23.5% fracture rate) and 468 (6.8%) from high to low risk (15.0% fracture rate) (eTable 3). McNemar’s test confirmed a significant difference in classification (χ² = 333.44, *p* < 0.001), indicating improved clinical stratification. Results were consistent with the SBayesR-based model.

### 3.5 Decision Curve Analysis (DCA)

Decision curve analyses demonstrated improved net clinical benefit for Bayes-FRAX compared to FRAX-CRF across clinically relevant MOF risk thresholds (10%-30%), notably at the commonly used 20% threshold. This analysis highlighted the superior clinical utility of Bayes-FRAX in guiding treatment decisions (Figure 3).

**Figure 3.**
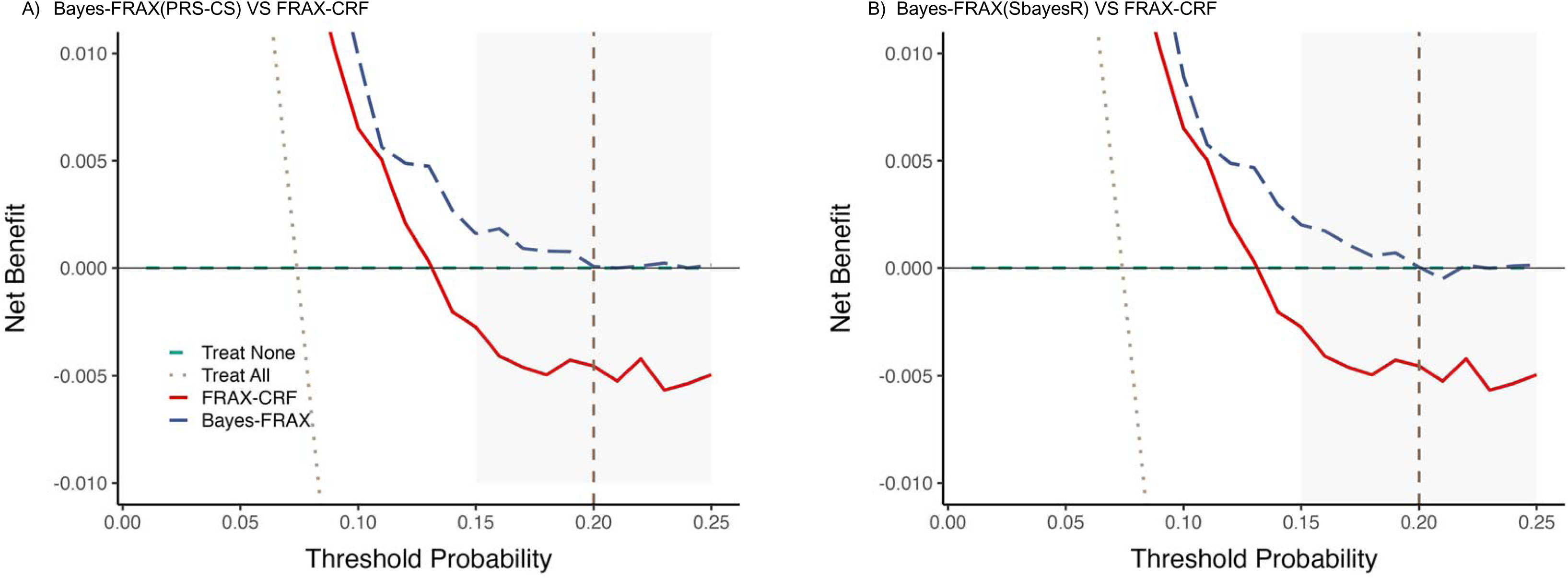
Decision curve analysis comparing the FRAX-CRF and Bayes-FRAX. The x-axis represents the threshold probability for starting treatment, and the y-axis represents the net benefit. Bayes-FRAX (dashed blue) consistently outperformed FRAX-CRF (solid red) between 10–20% thresholds, of fering greater benefit for guiding treatment decisions. The “treat all” line (dotted grey) assumes all individuals are treated, while the “treat none” (dashed green) assumes no one is treated. The dashed vertical line marks the 20% clinical cutoff.

### 3.6 Age-Stratified Analysis

Age-stratified analysis showed that fracture incidence remained higher in older individuals across all GPS quantiles, revealing the strong influence of age in combination with genetic risk. (eFigure 2). It confirmed that Bayes-FRAX had greater clinical utility in women >65 years. In this group, Bayes-FRAX showed improved AUROC (from 0.563 to 0.582) and net benefit across 5%–20 % risk thresholds. In contrast, improvements in younger women were minimal (eTable 4, eFigure 3).

### 3.7 Sensitivity Analysis Without BMD

Sensitivity analyses demonstrated consistent predictive performance of Bayes-FRAX with or without BMD. For both PRS-CS and SBayesR methods, AUROC remained stable when BMD was excluded (AUROC = 0.74 with BMD vs. 0.72 without BMD for both models; DeLong’s test *P* = 0.55 for PRS-CS and *P* = 0.59 for SBayesR). Sensitivity and specificity were 30.43% and 91.26% (PRS-CS) and 30.43% and 91.02% (SBayesR), respectively. NRI and integrated discrimination improvement (IDI) also remained robust without BMD (eTable 5).

### 3.8 External Validation

External validation performed in an independent cohort of 852 White postmenopausal women demonstrated comparable predictive performance. Odds ratios indicated significant associations with the risk of MOF (PRS-CS: OR=0.148, 95% CI: 0.052-0.411; SBayesR: OR=0.116, 95% CI: 0.040-0.324), confirming the robustness and generalizability of the Bayes-FRAX (eTable 6).

## Discussion

In this cohort study of postmenopausal women, integrating GPS derived using advanced Bayesian methods (PRS-CS and SBayesR) significantly improved fracture risk prediction accuracy compared to the standard FRAX with clinical risk factor (FRAX-CRF) only. Specifically, Bayes-FRAX demonstrated statistically significant enhancements in AUROC, AUPRC, calibration accuracy, and diagnostic sensitivity. These predictive improvements translated into measurable clinical reclassification benefits, particularly among women near established clinical intervention thresholds. Predictive enhancements were notably more pronounced in older women aged over 65 years, highlighting potential age-specific clinical applicability. External validation further confirmed the robustness and generalizability of these findings.

Previous PRS studies typically reported modest incremental improvements in fracture prediction beyond established clinical risk factors[18, 20]. A primary limitation of prior polygenic approaches has been their reliance on genome-wide significant variants, capturing only limited genetic susceptibility[21, 35]. In contrast, our Bayesian GPS approach explicitly accounts for linkage disequilibrium and complex polygenic architectures, thereby capturing broader genetic contributions to fracture susceptibility. Although absolute improvements in predictive accuracy were moderate (AUROC increased from 0.662 to approximately 0.680), these gains represent meaningful progress addressing well-documented predictive limitations of FRAX, particularly among individuals at or near clinical intervention thresholds.

Clinically, these predictive improvements may have practical implications by enhancing the accuracy of fracture risk stratification, particularly for patients near critical clinical decision thresholds. For instance, a 66-year-old woman with a FRAX-CRF score just below the 20% intervention threshold, who would typically not meet treatment criteria, might be reclassified to higher risk based on a high GPS indicating substantial genetic susceptibility. Conversely, a patient with a FRAX-CRF score slightly above the intervention threshold but possessing a low genetic risk score could potentially be considered for monitoring and lifestyle interventions instead of pharmacological treatment. These explicit clinical scenarios demonstrate how Bayes-FRAX models enhance shared clinical decision-making by providing nuanced, genetically informed risk assessments[36].

DCA explicitly demonstrated that Bayes-FRAX provides greater net clinical benefit than FRAX-CRF across clinically relevant MOF risk thresholds, particularly at the widely used 20% threshold. This highlights the explicit clinical utility of incorporating genetic risk scores in refining fracture risk estimates for individuals at intermediate risk levels, guiding more personalized and effective treatment decisions.

This study’s strengths include its large, well-characterized postmenopausal cohort, explicit integration of advanced Bayesian-derived GPS methods, rigorous internal cross-validation, external validation, and comprehensive clinical reclassification analysis. These methodological features collectively ensure the generalizability of the findings.

However, several limitations merit explicit acknowledgment. First, fracture outcomes relied heavily on self-reported data, which may be subject to recall or misclassification bias, though prior WHI validation studies indicate high specificity. Second, the predominantly White participant cohort limits the generalizability to ethnically diverse populations. Third, reliance on existing GWAS summary statistics inherently limits genetic variant coverage and predictive precision.

Future research directions should focus on validating Bayes-FRAX in ethnically diverse cohorts. Prospective studies or pilot clinical implementations should be conducted to assess real-world clinical utility, feasibility, and patient outcomes. Subsequently, economic evaluations should be conducted to evaluate whether the predictive improvement justifies the integration costs, including DNA sequencing, into routine osteoporosis management.

In conclusion, Bayes-FRAX explicitly demonstrates statistically significant enhancements in fracture risk prediction by integrating Bayesian-derived genome-wide polygenic scores into clinical risk assessment frameworks. While improvements are moderate, they suggest promising directions for personalized osteoporosis care. Further validation in diverse populations and practical clinical contexts remains essential before implementation in clinical settings.

## Data Availability

The individual-level data used in this study are available through controlled access from the database of Genotypes and Phenotypes (dbGaP) at https://www.ncbi.nlm.nih.gov/projects/ gap/cgi-bin/study.cgi?study_id=phs000200.v12.p3. The genome-wide association summary statistics were obtained from the GEnetic Factors for Osteoporosis (GEFOS) Consortium and are publicly available at http://www.gefos.org/?q=content/data-release-2018

## Author contributions

Conceptualization: Qing Wu. Formal analysis: Anqi Liu. Methodology: Qing Wu, Anqi Liu. Supervision: Qing Wu. Software: Anqi Liu. Writing –original draft: Anqi Liu, Jianing Liu, Qing Wu. Writing –review & editing: Jianing Liu, Qing Wu. Funding acquisition: Qing Wu. All authors have read and agreed to the published version of the manuscript.

## Data Sharing Statement

The individual-level data used in this study are available through controlled access from the database of Genotypes and Phenotypes (dbGaP) at https://www.ncbi.nlm.nih.gov/projects/gap/cgi-bin/study.cgi?study_id=phs000200.v12.p3. The genome-wide association summary statistics were obtained from the GEnetic Factors for Osteoporosis (GEFOS) Consortium and are publicly available at http://www.gefos.org/?q=content/data-release-2018

