## Supplemental Document for "Bayesian Genome-wide Polygenic Score Integration with FRAX for Enhanced Fracture Risk Prediction in Postmenopausal Women"

**Supplemental Content**

**eMethods 1. Prior Settings for Bayesian GPS Derivation Using SbayesR**

**eFigure 1.** Comparison of the predictive performance of FRAX-CRF and two Bayes-FRAX**.**

**eFigure 2. Association between Genetic Polygenic Score (GPS) quantiles and observed fracture rates.**

**eFigure 3. Decision curve analysis (DCA) stratified by age (≤65 and >65 years) comparing FRAX-CRF with Bayes-FRAX models.**

**eTable 1. Model performance metrics**

**eTable 2.** Cross-validation of model discrimination

**eTable 3.** Risk Reclassification between Standard FRAX and Bayes-FRAX and corresponding Fracture Incidence

**eTable 4.** **Sensitivity and age-stratified subgroup analysis comparing discrimination (AUROC) and net reclassification improvement (NRI) for FRAX-CRF and Bayes-FRAX models in women aged ≤65 years and >65 years**

**eTable 5**. Sensitivity Analysis of Predictive Performance of Bayes-FRAX model Models with and without BMD

**eTable 6.** Association between GPS and major osteoporotic fracture (MOF) based on logistic regression for external validation in independent cohort with 852 White participants.

**eMethods 1. Prior Settings for Bayesian GPS Derivation Using SbayesR**

**SBayesR assumes a mixture of normal distributions for SNP effects, controlled by two key priors:**

- **π: the proportion of SNPs in each mixture component**
- **γ: the corresponding variance for each component**

**The following combinations were used:**

| **GPS ID** | **π Settings** | **γ Settings** |
| --- | --- | --- |
| **1** | **0.50, 0.25, 0.15, 0.10** | **0.0, 0.001, 0.01, 0.1** |
| **2** | **0.10, 0.30, 0.30, 0.30** | **0.0, 0.001, 0.01, 0.1** |
| **3** | **0.01, 0.19, 0.30, 0.50** | **0.0, 0.001, 0.01, 0.1** |
| **4** | **0.05, 0.15, 0.30, 0.50** | **0.0, 0.001, 0.01, 0.1** |
| **5** | **0.10, 0.20, 0.30, 0.40** | **0.0, 0.001, 0.01, 0.1** |

**Supplementary Figure**

**eFigure 1. Comparison of the predictive performance of FRAX-CRF and two Bayes-FRAX.** ROC curves in panels (A) and (B) compare FRAX-CRF with Bayes-FRAX constructed using PRS-CS and SBayesR methods, respectively.

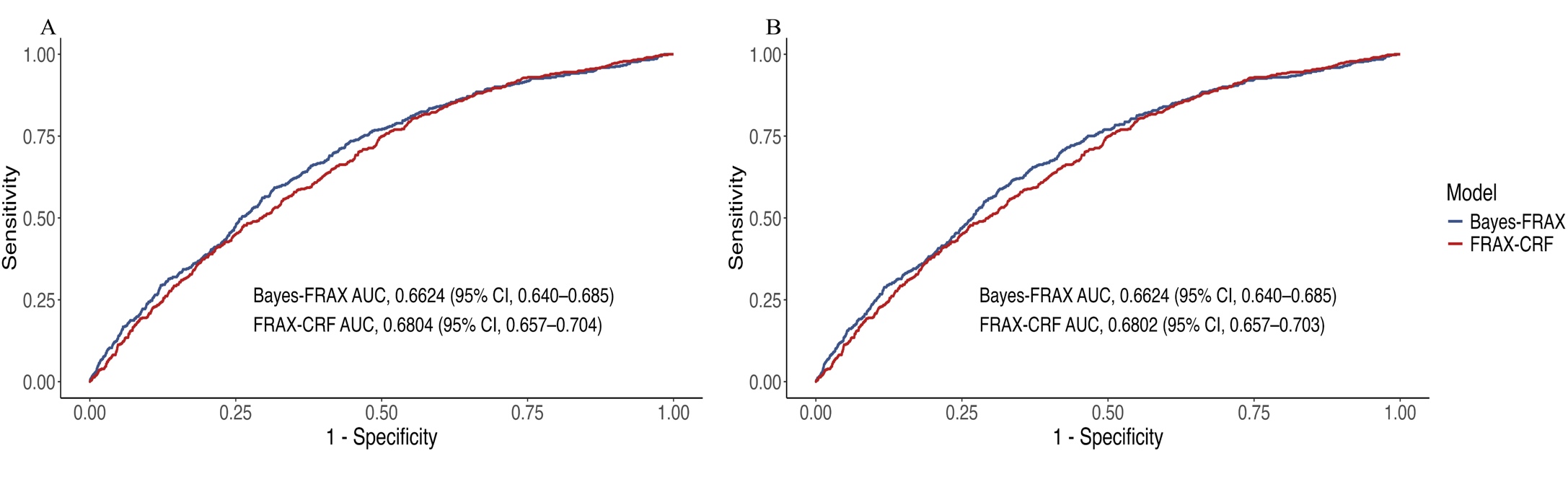

**eFigure 2. Association between Genetic Polygenic Score (GPS) quantiles and observed fracture rates. Lower GPS quantiles indicate higher genetic risk and are associated with a higher observed rate of major osteoporotic fractures. (A) Observed fracture rates stratified by GPS quantiles across age groups using PRS-CS-derived GPS. (B) Observed fracture rates stratified by GPS quantiles across age groups using SBayesR-derived GPS.**

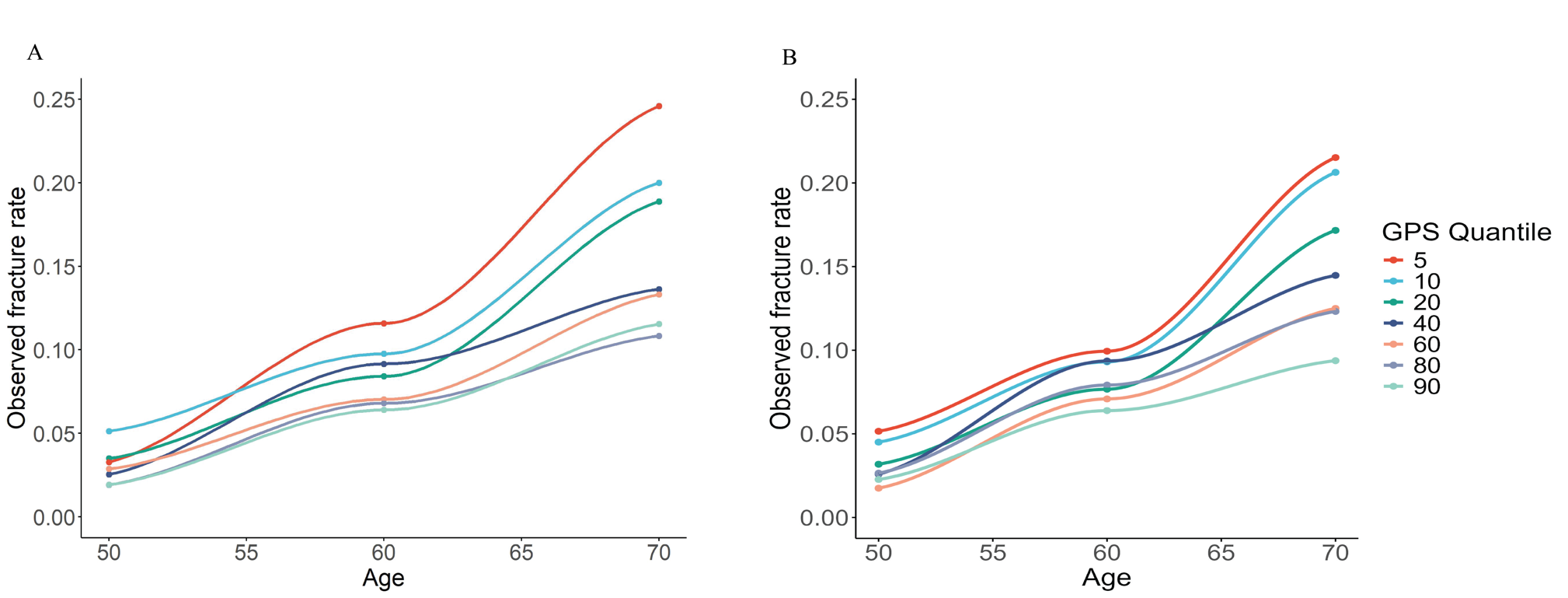

**eFigure 3,Decision curve analysis (DCA) stratified by age (≤65 and >65 years) comparing FRAX-CRF with Bayes-FRAX models.(A) Comparison between Bayes(PRS-CS)-FRAX and FRAX-CRF for participants aged ≤65 years.(B) Comparison between Bayes(SbayesR)-FRAX and FRAX-CRF for participants aged ≤65 years.(C) Comparison between Bayes(PRS-CS)-FRAX and FRAX-CRF for participants aged >65 years.(D) Comparison between Bayes(SbayesR)-FRAX and FRAX-CRF for participants aged >65 years. Dashed blue lines represent Bayes-FRAX models, solid red lines represent the FRAX-CRF model. The vertical dashed line marks the 20% intervention threshold.**

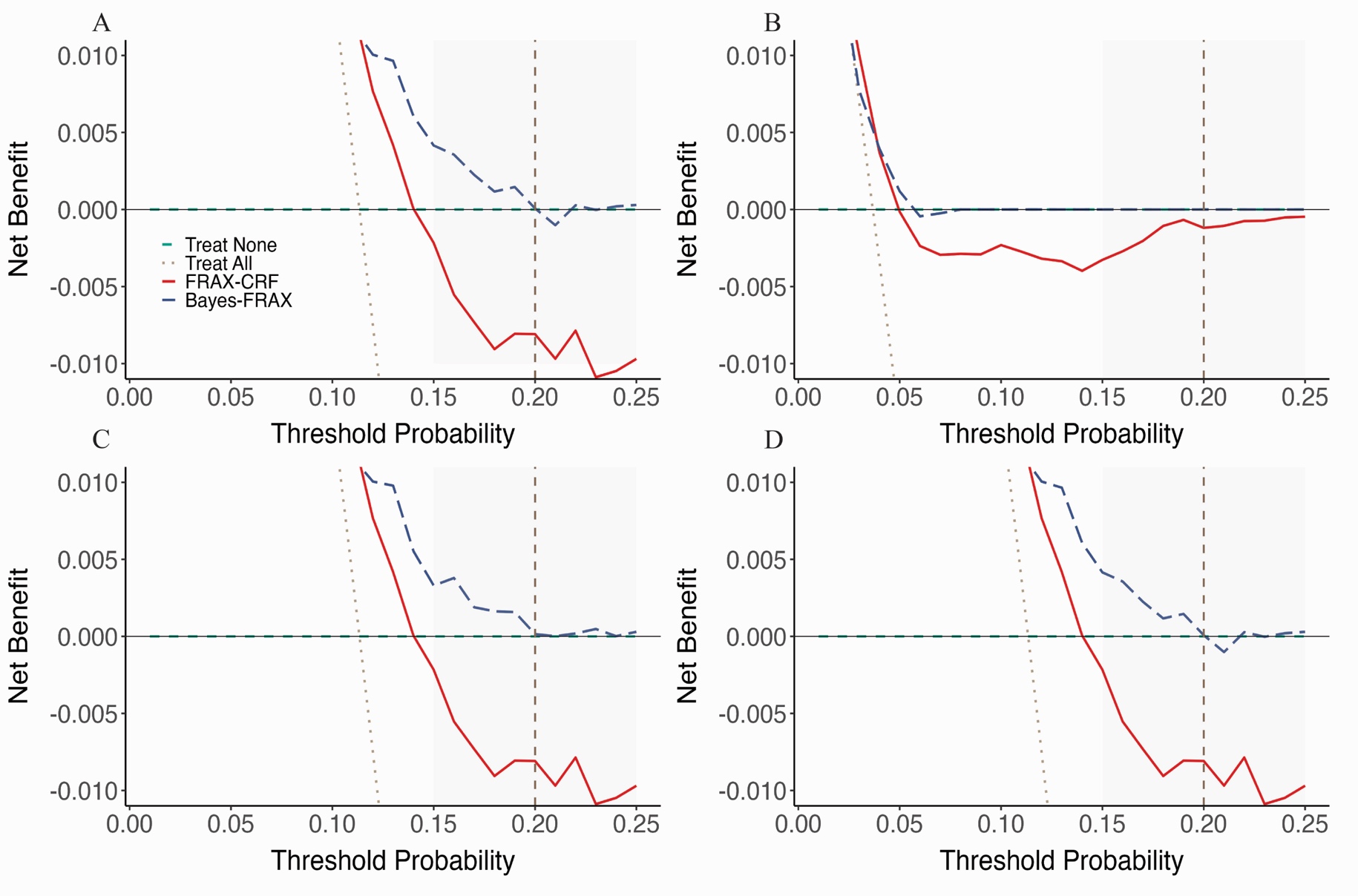

**Supplementary Table**

**eTable 1. Model performance metrics.** Summary of model discrimination, calibration, and reclassification performance for FRAX-CRF and Bayes-FRAX models. The Bayes-FRAX models incorporate genome-wide polygenic scores derived using PRS-CS and SBayesR methods. Metrics include AUROC (discrimination), AUPRC (precision-recall performance), NRI (reclassification), calibration slope with 95% confidence intervals, and Hosmer–Lemeshow goodness-of-fit p-values.

| Metric | FRAX-CRF | Bayes-FRAX(PRS-CS) | Bayes-FRAX(SbayesR) |
| --- | --- | --- | --- |
| AUROC | 0.6624(0.6396 – 0.6852) | 0.6804(0.6573-0.7035)^a^ | 0.6802(0.6571-0.7033)^b^ |
| AUPRC | 0.1198 | 0.1400 | 0.1380 |
| NRI | Reference | 4.3444% | 4.5861% |
| Calibration Slope (95% CI) | 1 (0.8168 - 1.1810) | 1 (0.8569 - 1.1431) | 1 (0.8560 - 1.1437) |
| Hosmer-Lemeshow p value | <0.0001 | 0.0013 | 0.0006 |

**^a^P = 0.045 vs FRAX-CRF (DeLong’s test)**

**^b^P =0.047 vs FRAX-CRF (DeLong’s test)**

**eTable 2.** **Cross-validation of model discrimination.** Area under the receiver operating characteristic curve (AUROC) values for the FRAX-CRF model and Bayes-FRAX models using polygenic scores derived via SBayesR and PRS-CS. Results are shown across five folds and as an overall mean from 5-fold cross-validation.

| Fold | AUROC(FRAX-CRF) | AUROC(SbayesR) | AUROC(PRS-CS) |
| --- | --- | --- | --- |
| 1 | 0.654 | 0.695 | 0.675 |
| 2 | 0.684 | 0.660 | 0.669 |
| 3 | 0.672 | 0.659 | 0.685 |
| 4 | 0.655 | 0.718 | 0.666 |
| 5 | 0.648 | 0.670 | 0.707 |
| Mean | 0.663 | 0.680 | 0.681 |

**eTable 3. Risk Reclassification between Standard FRAX and Bayes-FRAX and corresponding Fracture Incidence**

| Standard FRAX | Bayes-FRAX | N(PRS-CS) | Fracture (%) | N (SbayesR) | Fracture (%) |
| --- | --- | --- | --- | --- | --- |
| High | High | 53 | 9 (17.0%) | 49 | 8 (16.3%) |
| High | Low | 468 | 70 (15.0%) | 472 | 71 (15.0%) |
| Low | High | 51 | 12 (23.5%) | 49 | 12 (24.5%) |
| Low | Low | 6360 | 422 (6.6%) | 6362 | 422 (6.6%) |

**eTable 4.** **Sensitivity and age-stratified subgroup analysis** **comparing discrimination (AUROC) and net reclassification improvement (NRI) for FRAX-CRF and Bayes-FRAX models in women aged ≤65 years and >65 years. Results are presented separately for fixed intervention threshold (MOF risk ≥20%) and age-dependent thresholds (based on NOGG guidelines).**

|  | Age ≤65 | | | Age>65 | | |
| --- | --- | --- | --- | --- | --- | --- |
|  | AUROC | NRI  (Fixed MOF 20%) | NRI  (Age-dependent MOF) | AUROC | NRI  (Fixed MOF 20%) | NRI  (Age-dependent MOF) |
| FRAX-CRF | 0.595 | Reference | Reference | 0.563 | Reference | Reference |
| Bayes-FRAX(PRS-CS) | 0.592 | 0.084% | -0.122% | 0.582 | 4.679% | 4.585% |
| Bayes-FRAX(SbayesR) | 0.591 | 0.087% | -0.122% | 0.581 | 5.041% | 4.618% |

Abbreviations: MOF, Major Osteoporotic Fracture ; NOGG, National Osteoporosis Guideline Group

**eTable 5**. **Sensitivity Analysis of Predictive Performance of Bayes-FRAX model Models with and without BMD**

| Performance Metric | Bayes-FRAX(PRS-CS) (With BMD) | Bayes-FRAX(PRS-CS) (Without BMD) | DeLong’s test | Bayes-FRAX(SbayesR) (With BMD) | | Bayes-FRAX(SbayesR) (Without BMD) | DeLong’s test |
| --- | --- | --- | --- | --- | --- | --- | --- |
| AUROC | 0.74 | 0.72 | P = 0.55 | | 0.74 | 0.72 | P = 0.59 |
| Sensitivity (%) | 30.43 | 10.87 |  | | 30.43 | 10.87 |  |
| Specificity (%) | 91.26 | 91.50 |  | | 91.02 | 91.50 |  |
| NRI | 0.19 |  |  | | 0.19 |  |  |
| NRI (cases) | 0.20 |  |  | | 0.20 |  |  |
| NRI (controls) | 0.00 |  |  | | -0.01 |  |  |
| IDI | 0.03 |  |  | | 0.03 |  |  |

Abbreviations: IDI, Integrated Discrimination Improvement

**eTable 6.** **Association between GPS and major osteoporotic fracture (MOF) based on logistic regression for external validation in independent cohort with 852 White participants.** The table shows odds ratios (OR) and 95% confidence intervals (CI) for FRAX-CRF alone and when combined with genome-wide polygenic scores (GPS) derived using PRS-CS and SBayesR methods. Likelihood ratio tests compare the fit of models with and without GPS.

| Model |  | Odds ratio(95%CI) | P value |
| --- | --- | --- | --- |
| Fracture ~ FRAX-CRF | FRAX-CRF |  |  |
| Fracture~ FRAX-CRF+ GPS(PRS-CS) | FRAX-CRF | 1.075(1.052,1.100) |  |
|  | GPS(PRS-CS) | 0.148(0.052,0.411) | <0.001 |
| Likelihood ratio test |  |  |  |
| Fracture~FRAX-CRF | FRAX-CRF | 1.074(1.051,1.098) |  |
| Fracture~FRAX-CRF+GPS(SbayesR) | FRAX-CRF | 1.075(1.052,1.100) |  |
|  | GPS(SbayesR) | 0.116(0.040,0.324) |  |
| Likelihood ratio test |  |  | <0.001 |
